# Does Percutaneous Coronary Intervention Improve Survival for Out-Of-Hospital Cardiac Arrest Patients Receiving Extracorporeal Cardiopulmonary Resuscitation?

**DOI:** 10.64898/2026.07.27.26359075

**Authors:** Jake Toy, Kelsey Thompson, Nichole Bosson, Amir Abolhoda, Eddy Fan, Vadim Gudzenko, David Shavelle

## Abstract

**Introduction:** Extracorporeal cardiopulmonary resuscitation (ECPR) can support patients who fail to respond to standard resuscitation for out-of-hospital cardiac arrest (OHCA) allowing further time for critical interventions and patient recovery. Although the majority of patients with refractory OHCA have coronary artery disease, the role of emergent percutaneous coronary intervention (PCI) is not clear. We evaluated the effect of PCI on survival to hospital discharge (SHD) in a contemporary cohort of patients with OHCA receiving ECPR.

**Methods:** We performed a retrospective study using data from the Extracorporeal Life Support Organization (ELSO) registry. We included patients ≥18 years with OHCA due to a presumed cardiac etiology or an initial shockable rhythm who received ECPR from January 2020 to December 2023. Our primary outcome was SHD. We used inverse probability weighted matching to estimate the average treatment effect of PCI on SHD. We also performed a sensitivity analysis of patients most likely to benefit from PCI (witnessed arrest and no return of spontaneous circulation after five minutes of cardiopulmonary resuscitation.

**Results:** Of 1336 OHCA patients receiving ECPR, 1131 were included in the final analysis after exclusions for age (n=31) and presumed non-cardiac or initial non-shockable rhythm (n=174). The median age was 55 years (IQR 44-62) and most patients were male (n=901, 80%). Twenty-one percent (n=243) received PCI; those patients who received PCI were slightly older (58 [IQR 48-63] vs 53 [IQR 42-62]) and more often male (n=212 [87%] vs n=689 [78%]). In the primary analysis, we found no significant difference in SHD for patients who received PCI compared to those who did not receive PCI (–3.56%, 95% CI –10.31 to 3.19; p-value 0.301). In our sensitivity analysis, we also did not find a significant difference in SHD for patients who received PCI compared to those who did not receive PCI (–4.52%, 95% CI – 12.42 to 2.46; p-value 0.246).

**Conclusion:** In our registry-based study of refractory OHCA patients receiving ECPR, emergent PCI was not associated with a significant improvement in SHD.

**CLINICAL PERSPECTIVE:** *What Is New?:*

- This is the largest cohort to date of patients with out-of-hospital cardiac arrest undergoing extracorporeal cardiopulmonary resuscitation and percutaneous coronary intervention; this study evaluates the effect of percutaneous coronary intervention on survival to hospital discharge.

*2) What Are the Clinical Implications?:*

- The lack of improvement in survival suggests that additional prospective studies evaluating the timing of percutaneous coronary intervention are needed to further clarify its role within the extracorporeal cardiopulmonary resuscitation care pathway.

## INTRODUCTION

Extracorporeal cardiopulmonary resuscitation (ECPR) can be used as advanced therapy for patients who fail to respond to standard resuscitation for out-of-hospital cardiac arrest (OHCA) (1–3). The timing of ECPR, location of cannulation and patient selection continue to evolve (4–6). The goal of ECPR is to support the patients oxygenation and perfusion to allow for key interventions and patient recovery.

A significant proportion of patients with OHCA secondary to a shockable rhythm have acute coronary lesions and, therefore, may benefit from timely percutaneous coronary intervention (PCI) (7). Retrospective studies have failed to show a clear benefit of PCI in this patient population (8). Randomized studies evaluating the role of ECPR for refractory OHCA have yielded conflicting results and details regarding the benefits, or lack thereof for PCI, have not been explored (7, 9, 10).

We therefore sought to evaluate the effect of PCI on survival to hospital discharge (SHD) in a contemporary cohort of patients with OHCA receiving ECPR within the Extracorporeal Life Support Organization (ELSO) registry.

## METHODS

We performed a retrospective registry-based study to quantify the effect of PCI on survival to hospital discharge for OHCA patients receiving ECPR within the ELSO Registry. Our findings are reported in accordance with the Strengthening the Reporting of Observational Studies in Epidemiology (STROBE) guidelines (11). This study was determined to be exempt from human subjects review by the Institutional Review Board at the University of California, Los Angeles.

### Data Acquisition

We obtained data from the ELSO Registry, which is an international voluntary registry that collects information on adult and pediatric patients receiving ECMO. The ELSO Registry was established in 1989 and includes patients treated at over 500 hospitals across the globe. Data from the ELSO Registry has been widely used for quality improvement, regulatory approvals, and research. The structure of the registry and data collection methods have previously been described (12).

### Study Population

We included adult patients (age ≥18 years) with OHCA due to a presumed cardiac etiology or with an initial shockable rhythm who received ECPR from January 1, 2020 to December 31, 2023. We excluded patients >75 years old and those with a non-shockable rhythm and non-cardiac etiology for the arrest. These criteria were designed to largely align with prior clinical trials evaluating the impact of ECPR for OHCA (9, 10).

### Data Variables

We extracted data from the ELSO Registry including demographic and clinical variables. Demographic characteristics included age and sex. Clinical characteristics included location of arrest, provision of bystander cardiopulmonary resuscitation (CPR), bystander automated external defibrillator (AED) application, arrest witnessed status, initial rhythm, number of epinephrine doses administered, number of defibrillations or cardioversions, total CPR time, location of cannulation, pulse at time of cannulation, rearrest events prior to cannulation, rhythm at time of cannulation, PCI performed, and survival outcomes. Age, number of epinephrine doses, and number of defibrillations or cardioversions into quantiles were stratified and these were treated as polynomial variables. Current procedural terminology (CPT) codes were used to identify patients who received PCI (92920, 92921, 92924, 92925, 92928, 92929, 92933, 92934, 92937, 92941, 92943, 92944) and all PCI procedures occurred after ECPR initiation.

There was missingness among several covariates which we addressed through multiple imputations by chained equations (MICE) (13). We created 20 complete datasets using predictive mean matching for continuous variables, logistic regression for binary variables, and classification and regression trees for polynomial variables. We used the outcome, exposure, and all covariates for imputation of missing values. We imputed low missingness variables first followed by high missingness variables to improve imputation accuracy among high missingness variables.

### Outcome Measures

We defined SHD as our primary outcome. We treated SHD as a binary outcome at hospital discharge from the index hospitalization.

### Statistical Analysis

For descriptive statistics, we reported continuous variables as medians and interquartile ranges (IQR) and categorical variables as frequencies and percentages. We conducted an *a priori* sample size calculation based on an 18% survival to hospital discharge rate as described in a prior study of ECPR patients from the ELSO Registry (8). To detect an absolute difference of 10% in SHD as a clinically meaningful difference associated with PCI as estimated from prior literature (14), we chose a power of 80% and two-sided alpha of 0.05. Under this assumption, we estimated we would need at least 277 patients in each group.

We evaluated the primary outcome using a causal inference approach to estimate the average treatment effect (ATE) in our sample population. We selected ATE as our target estimate to understand the effect of PCI if performed on all patients with OHCA receiving ECPR. We adjusted for baseline confounders using inverse probability weighting (IPW) and assumed the time of treatment assignment (time zero) to be the time of cannulation. We selected baseline demographic and clinical covariates know at the hypothetical time zero, which could plausibly be associated with the intervention and outcome; these included year of incident, age, sex, location of arrest, provision of bystander CPR or AED use, arrest witnessed status, initial rhythm, number of epinephrine and defibrillations or cardioversions, and total CPR time. We estimated the relationship between these predictors and the intervention (e.g., PCI) using a generalized linear model (GLM) (Supplement A). Using a doubly robust approach, we estimated patient-level IPWs with a GLM using a logit link function within each imputed dataset and then fit a weighted generalized linear model in to estimate the treatment effect of PCI on SHD.^15^ We stabilized weights and trimmed extreme weights. The balance between baseline covariates before and after application of IPWs was assessed using standard mean differences and values <0.05 were considered adequately balanced (Supplement B). We reported 95% confidence intervals (CI) and considered p-values <0.05 as significant. We performed all analyses in R (Version 5.4.1, R Foundation for Statistical Computing, Vienna, Austria).

### Sensitivity Analysis

We defined one *a priori* sensitivity analysis, which was designed to isolate OHCA patients receiving ECPR that were most likely to benefit from PCI and with criteria that more closely aligned with recent ECPR trials (9, 10). In addition to age (18-75 years) and presumed cardiac etiology or initial shockable rhythm, we further selected patients with a witnessed arrest and no return of spontaneous circulation (ROSC) after five minutes of CPR. We also excluded patients who received ECPR for ≤1 hour and those with ECPR that was discontinued due to death or poor prognosis as these patients are unlikely to receive PCI in clinical practice. We repeated the complete analysis for this sensitivity.

## RESULTS

Of 1336 OHCA patients receiving ECPR, 1131 were included in the final analysis after exclusions for age (n=31) and presumed non-cardiac or initial non-shockable rhythm (n=174) (Figure 1). The median (IQR) age was 55 (44-62) years and the majority of patients were male (n=901, 80%). Twenty-one percent (n=243) received PCI; those patients who received PCI were slightly older (58 [IQR 48-63] vs 53 [IQR 42-62]) and more often male (n=212 [87%] vs n=689 [78%]). In the primary analysis, we found no significant difference in SHD for patients who received PCI compared to those who did not receive PCI (–3.56%, 95% CI –10.31 to 3.19; p-value 0.301). In our sensitivity analysis, we also did not find a significant difference in SHD for patients who received PCI compared to those who did not receive PCI (–4.52%, 95% CI –12.42 to 2.46; p-value 0.246).

**Figure 1.**
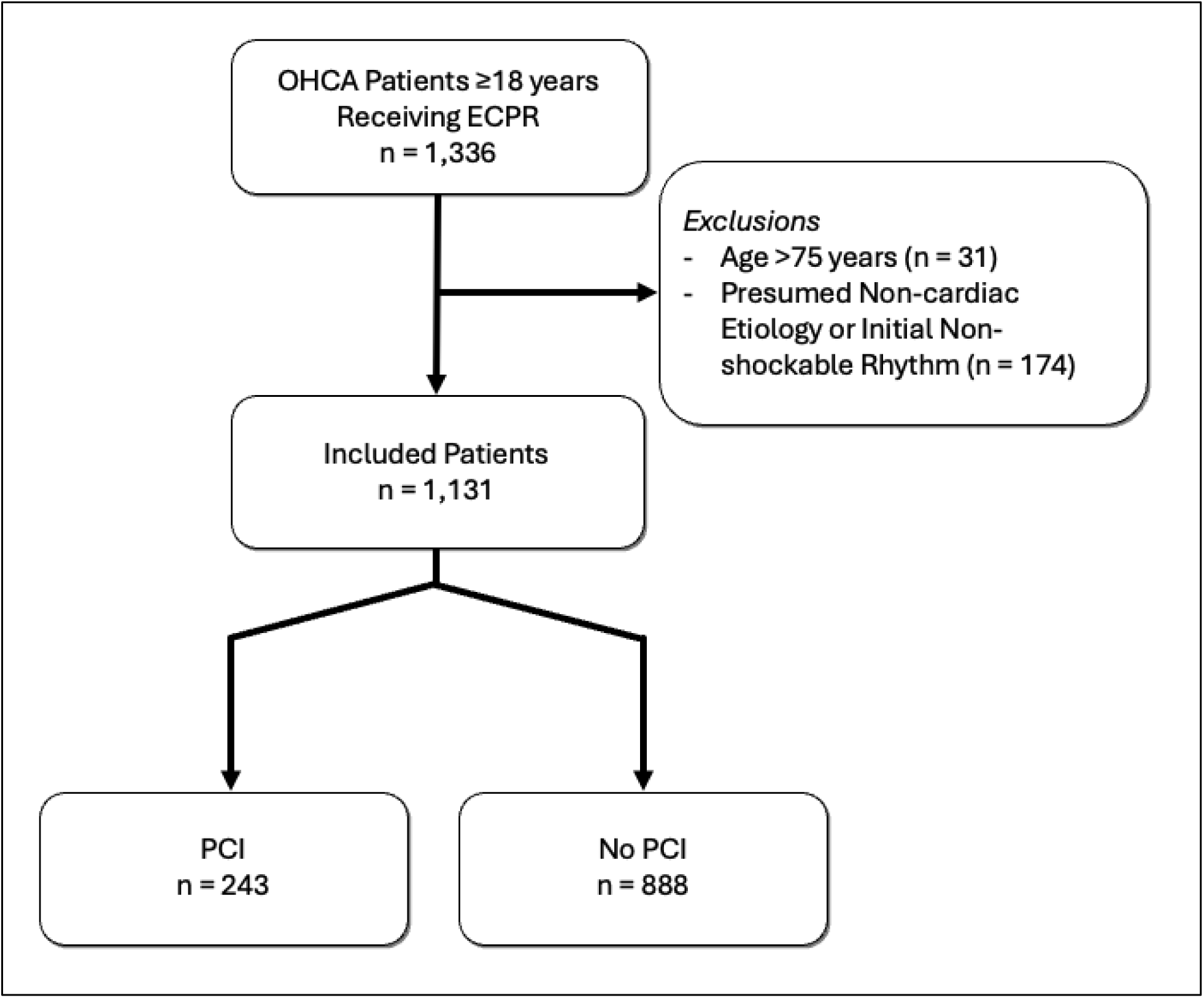
Study flow diagram. OHCA = out-of-hospital cardiac arrest, ECPR = extracorporeal cardiopulmonary resuscitation, PCI = percutaneous coronary intervention.

## DISCUSSION

In this investigation of a large international ECPR registry, we did not find a significant association between PCI and SHD among patients with OHCA who received ECPR. These findings were robust in our pre-specified sensitivity analysis, which aimed to create an enriched sample of refractory OHCA patients who received ECPR and were more likely to benefit from PCI. Our findings are consistent with prior retrospective analyses and randomized trials that evaluated the effect of PCI in refractory OHCA patients receiving ECPR (7, 8, 10).

These findings are similar to those from the study by Alhuneafat and colleagues (8). These investigators similarly applied a causal inference framework using ELSO Registry data and found no significant survival benefit associated with PCI following ECPR for OHCA. Notably, there are important population-level and methodological differences that distinguish these studies. Our analysis incorporated a substantially larger patient cohort who received PCI (243 versus 138 patients). Additionally, we applied a more stringent exclusion criteria to isolate those that may be more likely to benefit from PCI which more closely aligns with prior randomized trials (7, 9, 10) and prehospital ECPR routing guidelines (16). Our study excluded patients >75 years, those with an unwitnessed cardiac arrest but also excluded those with any ROSC within five minutes of CPR initiation and those with resuscitation after less than one hour of ECPR, two groups of patients less likely to benefit from an early PCI approach. In contrast, Alhuneafat et al. applied fewer exclusion criteria, resulting in a more heterogeneous study population, which could have diluted any effect (8). However, the consistent null effect seen in the current analysis supports their findings and suggests that PCI may not independently improve survival in OHCA patients receiving ECPR.

Our results should be interpreted in the context of the broader literature on coronary angiography and PCI in OHCA. Multiple analyses have demonstrated that early coronary angiography (CAG) is associated with improved survival in select OHCA populations, particularly those with ST-elevation myocardial infarction (STEMI), in whom the rate of acute coronary occlusion is substantially higher than with non-STEMI (16). However, generalized use of early CAG across all OHCA patients has not been shown to confer a survival benefit or improve neurological outcomes (17). Current guidelines nonetheless recognize specific indications for CAG in OHCA beyond STEMI, including refractory ventricular tachycardia or fibrillation and recurrent hemodynamic instability, which overlap substantially with the clinical indications driving ECPR use (1). This intersection is relevant as recurrent OHCA has been associated with more severe coronary artery disease burden (18), and among OHCA patients placed on ECPR who underwent immediate coronary angiography, a majority were found to have multivessel disease (16). Prior studies evaluating the use of PCI in patients with OHCA and receiving ECPR were limited by small sample size (7–10). Our study represents the largest dedicated analysis of PCI outcomes in ECPR-supported patients with OHCA to date and contributes important evidence where equipoise remains.

An important question, which may impact the effect of PCI for patients receiving ECPR, is procedural timing. Alhuneafat and colleagues included a stratified analysis for early (≤24 hours) or late (>24 hours) PCI procedural timing, which did not show a significant difference in the impact on survival (8). While all PCI occurred during ECPR in the current study, more detailed information about the exact timing of PCI after ECPR initiation was not an available data element in our study dataset. We were therefore unable to evaluate the potential effect of PCI timing on outcome. This creates the potential for immortal time bias. This is a significant limitation in the current analysis given that time-sensitive associations have been identified in this population which highlight a need for understanding the impact of timing of ECPR and PCI performance both as bundled ECPR procedures and in post arrest management in general. Longer time to ECPR initiation has been associated with lower rates of PCI performance, but earlier PCI, particularly within the first hour following initial collapse, has been associated with substantially improved survival (14, 19) consistent with the established benefit of early reperfusion in acute coronary syndromes (20). Given those findings, it is surprising that bundled resuscitation strategy for OHCA incorporating rapid intra-arrest transport, ECPR, and immediate invasive assessment did not demonstrate improved survival rates in the Prague OHCA trial, suggesting that procedural integration alone may be insufficient without careful patient selection (10). Recent randomized trials in patients with OHCA *not* undergoing ECPR and without ST segment elevation have failed to show a benefit of immediate angiography compared with delayed angiography (21, 22). As such, clear indications and timing of CAG in the OHCA population remain unclear. Present recommendations for ECPR on the basis of expert consensus includes both establishment of standardized post resuscitation protocols and interdisciplinary coordination and also recommend urgent CAG for all patients unless there is clear noncardiac cause of arrest (1). As the use of ECPR evolves, a more nuanced understanding of the timing of procedures will allow better optimization of post resuscitation protocols and multidisciplinary coordination. In addition, the timing of PCI for refractory OHCA receiving ECPR must be further explored in future prospective investigations.

## LIMITATIONS

Our study has several limitations. First, our study included a relatively small number of patients and may have been underpowered to detect a meaningful effect of PCI on survival. However, the ELSO registry is the largest contemporary database of ECPR patients and future analyses should be considered with the inclusion of subsequent years of data to increase the number of patients. Second, while we applied causal inference methodology, there is likely unmeasured confounding that may impact our ability to estimate the true effect of PCI. Third, this registry is subject to missingness; of particular concern is the potential missingness among CPT codes for PCI attached to each patient encounter. Fourth, our dataset did not include information on procedure timing which limited our ability to account for procedure timing in IPWs and creating a risk for immortal time bias. Lastly, there were no geographic or hospital indicators limiting our ability to account for region and/or site-related clustering.

## CONCLUSIONS

In our registry-based investigation of patients with refractory OHCA receiving ECPR, PCI was not associated with a significant improvement in SHD. This finding was consistent in both the primary analysis and the sensitivity analysis including patients most likely to benefit from coronary revascularization. Additional prospective studies evaluating PCI and the timing of this critical intervention are needed to further clarify the role of PCI within the ECPR care pathway.

## Data Availability

All of the data included in the manuscript is from the ELSO registry.

## ACKNOWLEDGEMENTS

We acknowledge the Extracorporeal Life Support Organization Registry team for assistance with data acquisition and study design.

## Nonstandard Abbreviations and Acronyms

ECPR: Extracorporeal cardiopulmonary resuscitation
OCHA: Out of hospital cardiac arrest
PCI: Percutaneous coronary intervention
ELSO: Extracorporeal life support organization
CPR: Cardiopulmonary resuscitation

## Tables and Figures

**Table 1:**
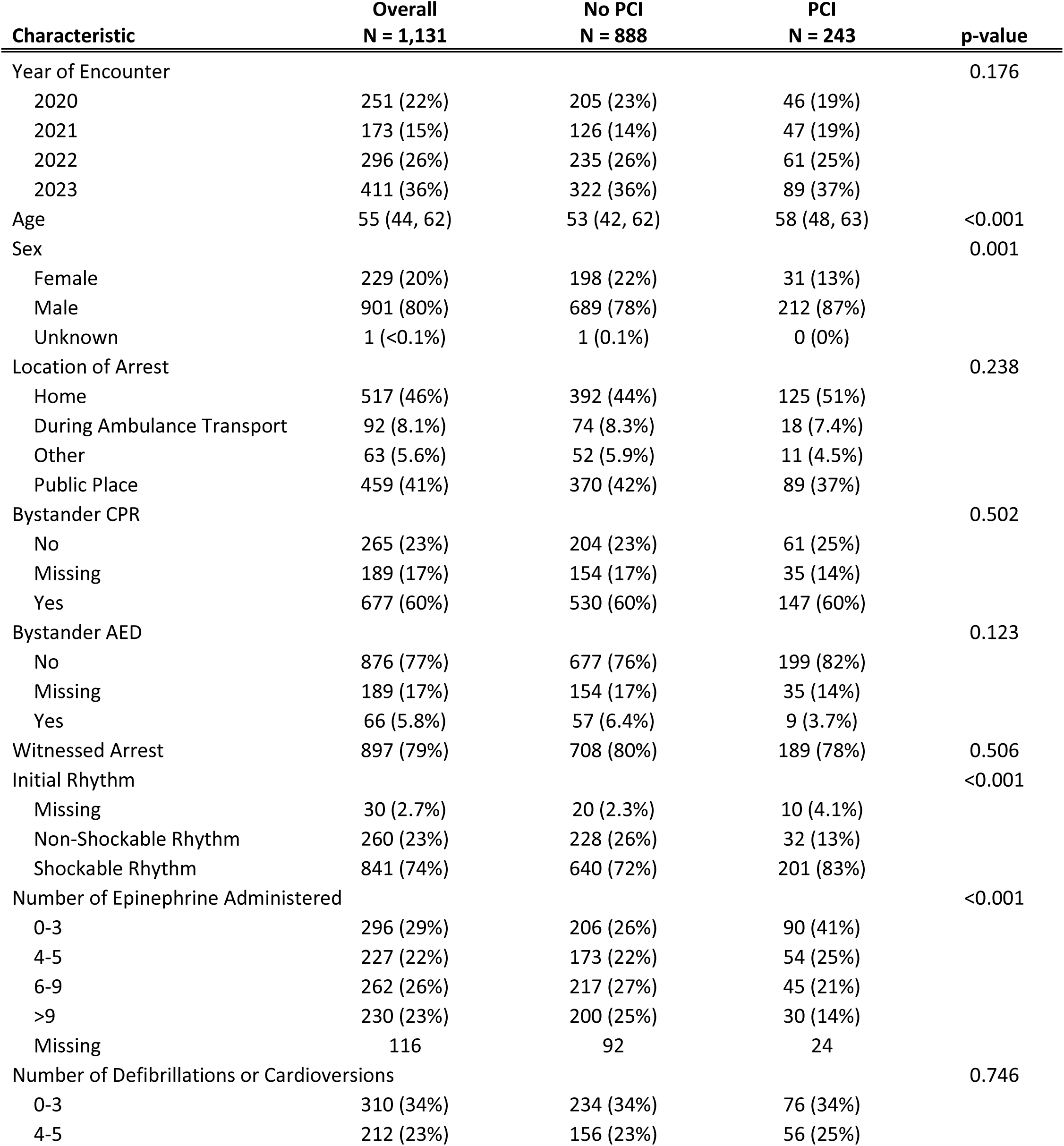
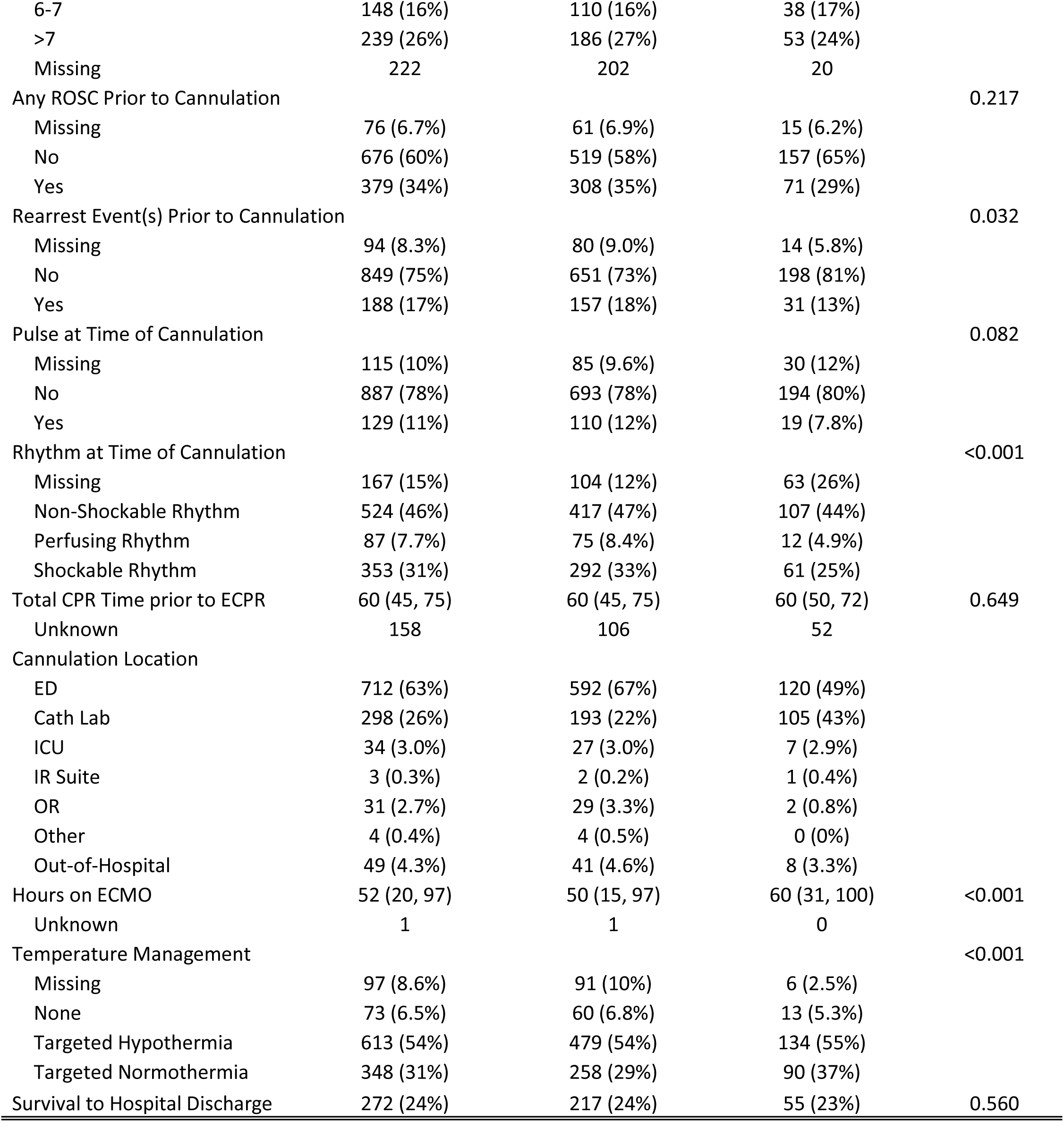
Demographic and clinical characteristics.

| Characteristic | Overall<br>N = 1,131 | No PCI<br>N = 888 | PCI<br>N = 243 | p-value |
| --- | --- | --- | --- | --- |
| Year of Encounter |  |  |  | 0.176 |
| 2020 | 251 (22%) | 205 (23%) | 46 (19%) |  |
| 2021 | 173 (15%) | 126 (14%) | 47 (19%) |  |
| 2022 | 296 (26%) | 235 (26%) | 61 (25%) |  |
| 2023 | 411 (36%) | 322 (36%) | 89 (37%) |  |
| Age | 55 (44, 62) | 53 (42, 62) | 58 (48, 63) | <0.001 |
| Sex |  |  |  | 0.001 |
| Female | 229 (20%) | 198 (22%) | 31 (13%) |  |
| Male | 901 (80%) | 689 (78%) | 212 (87%) |  |
| Unknown | 1 (<0.1%) | 1 (0.1%) | 0 (0%) |  |
| Location of Arrest |  |  |  | 0.238 |
| Home | 517 (46%) | 392 (44%) | 125 (51%) |  |
| During Ambulance Transport | 92 (8.1%) | 74 (8.3%) | 18 (7.4%) |  |
| Other | 63 (5.6%) | 52 (5.9%) | 11 (4.5%) |  |
| Public Place | 459 (41%) | 370 (42%) | 89 (37%) |  |
| Bystander CPR |  |  |  | 0.502 |
| No | 265 (23%) | 204 (23%) | 61 (25%) |  |
| Missing | 189 (17%) | 154 (17%) | 35 (14%) |  |
| Yes | 677 (60%) | 530 (60%) | 147 (60%) |  |
| Bystander AED |  |  |  | 0.123 |
| No | 876 (77%) | 677 (76%) | 199 (82%) |  |
| Missing | 189 (17%) | 154 (17%) | 35 (14%) |  |
| Yes | 66 (5.8%) | 57 (6.4%) | 9 (3.7%) |  |
| Witnessed Arrest | 897 (79%) | 708 (80%) | 189 (78%) | 0.506 |
| Initial Rhythm |  |  |  | <0.001 |
| Missing | 30 (2.7%) | 20 (2.3%) | 10 (4.1%) |  |
| Non-Shockable Rhythm | 260 (23%) | 228 (26%) | 32 (13%) |  |
| Shockable Rhythm | 841 (74%) | 640 (72%) | 201 (83%) |  |
| Number of Epinephrine Administered |  |  |  | <0.001 |
| 0-3 | 296 (29%) | 206 (26%) | 90 (41%) |  |
| 4-5 | 227 (22%) | 173 (22%) | 54 (25%) |  |
| 6-9 | 262 (26%) | 217 (27%) | 45 (21%) |  |
| >9 | 230 (23%) | 200 (25%) | 30 (14%) |  |
| Missing | 116 | 92 | 24 |  |
| Number of Defibrillations or Cardioversions |  |  |  | 0.746 |
| 0-3 | 310 (34%) | 234 (34%) | 76 (34%) |  |
| 4-5 | 212 (23%) | 156 (23%) | 56 (25%) |  |
| 6-7 | 148 (16%) | 110 (16%) | 38 (17%) |  |
| >7 | 239 (26%) | 186 (27%) | 53 (24%) |  |
| Missing | 222 | 202 | 20 |  |
| Any ROSC Prior to Cannulation |  |  |  | 0.217 |
| Missing | 76 (6.7%) | 61 (6.9%) | 15 (6.2%) |  |
| No | 676 (60%) | 519 (58%) | 157 (65%) |  |
| Yes | 379 (34%) | 308 (35%) | 71 (29%) |  |
| Rearrest Event(s) Prior to Cannulation |  |  |  | 0.032 |
| Missing | 94 (8.3%) | 80 (9.0%) | 14 (5.8%) |  |
| No | 849 (75%) | 651 (73%) | 198 (81%) |  |
| Yes | 188 (17%) | 157 (18%) | 31 (13%) |  |
| Pulse at Time of Cannulation |  |  |  | 0.082 |
| Missing | 115 (10%) | 85 (9.6%) | 30 (12%) |  |
| No | 887 (78%) | 693 (78%) | 194 (80%) |  |
| Yes | 129 (11%) | 110 (12%) | 19 (7.8%) |  |
| Rhythm at Time of Cannulation |  |  |  | <0.001 |
| Missing | 167 (15%) | 104 (12%) | 63 (26%) |  |
| Non-Shockable Rhythm | 524 (46%) | 417 (47%) | 107 (44%) |  |
| Perfusing Rhythm | 87 (7.7%) | 75 (8.4%) | 12 (4.9%) |  |
| Shockable Rhythm | 353 (31%) | 292 (33%) | 61 (25%) |  |
| Total CPR Time prior to ECPR | 60 (45, 75) | 60 (45, 75) | 60 (50, 72) | 0.649 |
| Unknown | 158 | 106 | 52 |  |
| Cannulation Location |  |  |  |  |
| ED | 712 (63%) | 592 (67%) | 120 (49%) |  |
| Cath Lab | 298 (26%) | 193 (22%) | 105 (43%) |  |
| ICU | 34 (3.0%) | 27 (3.0%) | 7 (2.9%) |  |
| IR Suite | 3 (0.3%) | 2 (0.2%) | 1 (0.4%) |  |
| OR | 31 (2.7%) | 29 (3.3%) | 2 (0.8%) |  |
| Other | 4 (0.4%) | 4 (0.5%) | 0 (0%) |  |
| Out-of-Hospital | 49 (4.3%) | 41 (4.6%) | 8 (3.3%) |  |
| Hours on ECMO | 52 (20, 97) | 50 (15, 97) | 60 (31, 100) | <0.001 |
| Unknown | 1 | 1 | 0 |  |
| Temperature Management |  |  |  | <0.001 |
| Missing | 97 (8.6%) | 91 (10%) | 6 (2.5%) |  |
| None | 73 (6.5%) | 60 (6.8%) | 13 (5.3%) |  |
| Targeted Hypothermia | 613 (54%) | 479 (54%) | 134 (55%) |  |
| Targeted Normothermia | 348 (31%) | 258 (29%) | 90 (37%) |  |
| Survival to Hospital Discharge | 272 (24%) | 217 (24%) | 55 (23%) | 0.560 |

## Supplemental Material

### Supplement A

**Table S1.** Predictors of percutaneous coronary intervention.

|  | Odds Ratio (CI) | P value |
| --- | --- | --- |
| Year of Encounter |  |  |
| 2021 | 1.73 (1.06, 2.82) | 0.029 |
| 2022 | 1.24 (0.79, 1.94) | 0.351 |
| 2023 | 1.19 (0.79, 1.81) | 0.407 |
| Age |  |  |
| 45-55 years | 2.27 (1.4, 3.67) | 0.001 |
| 56-63 years | 2.86 (1.79, 4.58) | 0 |
| 64-75 years | 2.39 (1.46, 3.9) | 0.001 |
| Male | 1.94 (1.25, 3) | 0.003 |
| Location of Arrest |  |  |
| During Ambulance Transport | 0.93 (0.49, 1.75) | 0.811 |
| Other | 0.84 (0.41, 1.74) | 0.643 |
| Public Place | 0.75 (0.54, 1.05) | 0.092 |
| Bystander CPR | 0.88 (0.59, 1.32) | 0.534 |
| AED | 0.66 (0.32, 1.35) | 0.259 |
| Witnessed Arrest | 0.94 (0.64, 1.36) | 0.733 |
| Initial Shockable Rhythm | 2.04 (1.32, 3.16) | 0.001 |
| Number of Epinephrine Administered |  |  |
| 4-5 | 0.8 (0.53, 1.22) | 0.307 |
| 6-9 | 0.51 (0.33, 0.8) | 0.004 |
| >9 | 0.37 (0.22, 0.6) | 0 |
| Number of Defibrillations or Cardioversions |  |  |
| 4-5 | 1.04 (0.68, 1.58) | 0.86 |
| 6-7 | 1.09 (0.68, 1.76) | 0.715 |
| >7 | 0.99 (0.63, 1.58) | 0.978 |
| Duration of CPR prior to ECMO | 1 (1, 1.01) | 0.327 |

**Figure S1.**
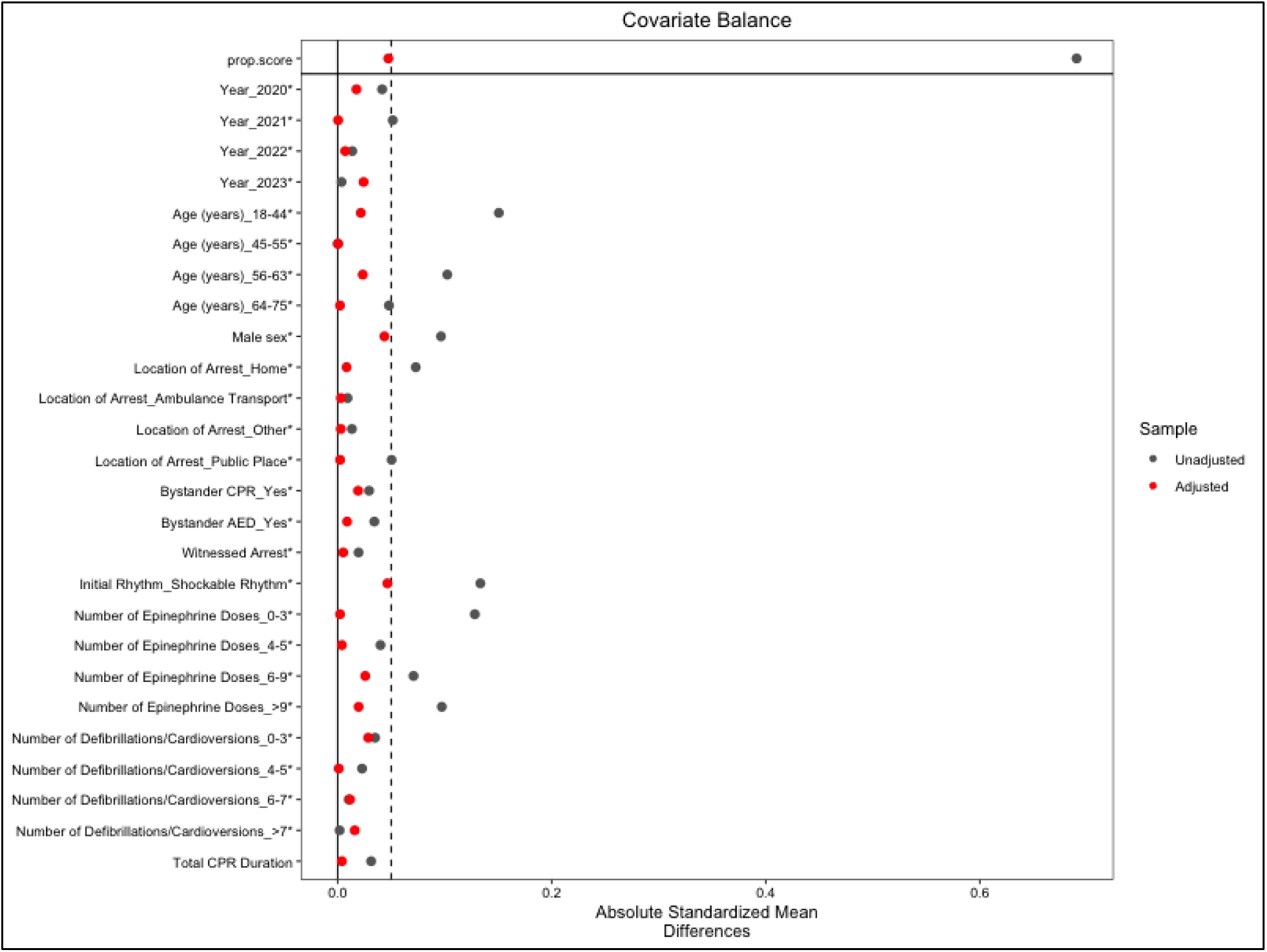
Love plot for covariate balance between group receiving percutaneous coronary intervention and not receiving percutaneous coronary intervention before and after weighting. CPR = cardiopulmonary resuscitation, AED = automated external defibrillator

### Supplement B

**Figure S2.**
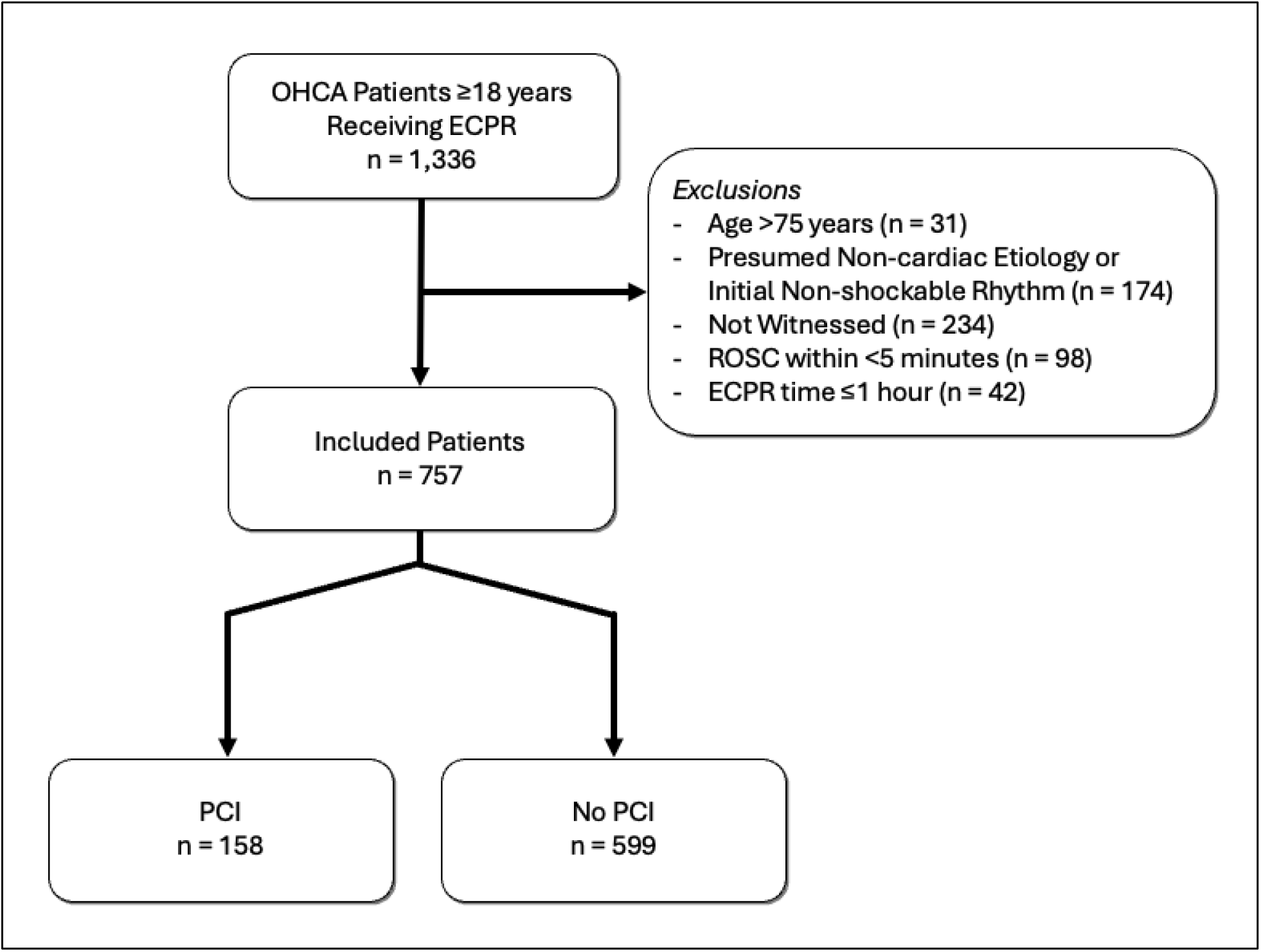
Sensitivity analysis flow diagram. OHCA = out-of-hospital cardiac arrest, ECPR = extracorporeal cardiopulmonary resuscitation, PCI = percutaneous coronary intervention, ROSC = return of spontaneous circulation

**Table S2.** Demographic and clinical characteristics for the sensitivity analysis.

| Characteristic | Overall<br>N = 757 | No PCI<br>N = 599 | PCI<br>N = 158 | p-value |
| --- | --- | --- | --- | --- |
| Year of Encounter |  |  |  | 0.688 |
| 2020 | 147 (19%) | 114 (19%) | 33 (21%) |  |
| 2021 | 107 (14%) | 81 (14%) | 26 (16%) |  |
| 2022 | 214 (28%) | 173 (29%) | 41 (26%) |  |
| 2023 | 289 (38%) | 231 (39%) | 58 (37%) |  |
| Age | 54 (45, 62) | 54 (43, 61) | 57 (48, 63) | 0.002 |
| Sex |  |  |  | 0.004 |
| Female | 152 (20%) | 133 (22%) | 19 (12%) |  |
| Male | 605 (80%) | 466 (78%) | 139 (88%) |  |
| Unknown | 0 (0%) | 0 (0%) | 0 (0%) |  |
| Location of Arrest |  |  |  | 0.828 |
| Home | 320 (42%) | 248 (41%) | 72 (46%) |  |
| During Ambulance Transport | 71 (9.4%) | 57 (9.5%) | 14 (8.9%) |  |
| Other | 46 (6.1%) | 37 (6.2%) | 9 (5.7%) |  |
| Public Place | 320 (42%) | 257 (43%) | 63 (40%) |  |
| Bystander CPR |  |  |  | 0.938 |
| No | 171 (23%) | 134 (22%) | 37 (23%) |  |
| Missing | 91 (12%) | 73 (12%) | 18 (11%) |  |
| Yes | 495 (65%) | 392 (65%) | 103 (65%) |  |
| Bystander AED |  |  |  | 0.363 |
| No | 614 (81%) | 481 (80%) | 133 (84%) |  |
| Missing | 91 (12%) | 73 (12%) | 18 (11%) |  |
| Yes | 52 (6.9%) | 45 (7.5%) | 7 (4.4%) |  |
| Initial Rhythm |  |  |  | 0.046 |
| Missing | 17 (2.2%) | 13 (2.2%) | 4 (2.5%) |  |
| Non-Shockable Rhythm | 174 (23%) | 149 (25%) | 25 (16%) |  |
| Shockable Rhythm | 566 (75%) | 437 (73%) | 129 (82%) |  |
| Precipitating Event |  |  |  | 0.032 |
| Cardiac | 673 (98%) | 522 (97%) | 151 (100%) |  |
| Non-Cardiac | 17 (2.5%) | 17 (3.2%) | 0 (0%) |  |
| Unknown | 67 | 60 | 7 |  |
| Number of Epinephrine Administered |  |  |  | 0.064 |
| 0-3 | 185 (27%) | 135 (25%) | 50 (34%) |  |
| 4-5 | 148 (22%) | 115 (21%) | 33 (23%) |  |
| 6-9 | 198 (29%) | 158 (29%) | 40 (27%) |  |
| >9 | 152 (22%) | 129 (24%) | 23 (16%) |  |
| Unknown | 74 | 62 | 12 |  |
| Number of Defibrillations or Cardioversions |  |  |  | 0.801 |
| 0-3 | 209 (33%) | 159 (33%) | 50 (35%) |  |
| 4-5 | 144 (23%) | 112 (23%) | 32 (22%) |  |
| 6-7 | 110 (18%) | 82 (17%) | 28 (19%) |  |
| >7 | 164 (26%) | 130 (27%) | 34 (24%) |  |
| Unknown | 130 | 116 | 14 |  |
| Any ROSC Prior to Cannulation |  |  |  | 0.010 |
| Missing | 28 (3.7%) | 19 (3.2%) | 9 (5.7%) |  |
| No | 460 (61%) | 352 (59%) | 108 (68%) |  |
| Yes | 269 (36%) | 228 (38%) | 41 (26%) |  |
| Rearrest Event(s) Prior to Cannulation |  |  |  | 0.264 |
| Missing | 40 (5.3%) | 32 (5.3%) | 8 (5.1%) |  |
| No | 589 (78%) | 459 (77%) | 130 (82%) |  |
| Yes | 128 (17%) | 108 (18%) | 20 (13%) |  |
| Pulse at Time of Cannulation |  |  |  | 0.026 |
| Missing | 47 (6.2%) | 31 (5.2%) | 16 (10%) |  |
| No | 619 (82%) | 490 (82%) | 129 (82%) |  |
| Yes | 91 (12%) | 78 (13%) | 13 (8.2%) |  |
| Rhythm at Time of Cannulation |  |  |  | 0.072 |
| Missing | 82 (11%) | 58 (9.7%) | 24 (15%) |  |
| Non-Shockable Rhythm | 354 (47%) | 276 (46%) | 78 (49%) |  |
| Perfusing Rhythm | 68 (9.0%) | 59 (9.8%) | 9 (5.7%) |  |
| Shockable Rhythm | 253 (33%) | 206 (34%) | 47 (30%) |  |
| Total CPR Time prior to ECPR | 58 (45, 73) | 57 (43, 74) | 60 (50, 73) | 0.204 |
| Cannulation Location |  |  |  |  |
| ED | 519 (69%) | 429 (72%) | 90 (57%) |  |
| Cath Lab | 146 (19%) | 92 (15%) | 54 (34%) |  |
| ICU | 29 (3.8%) | 22 (3.7%) | 7 (4.4%) |  |
| IR Suite | 3 (0.4%) | 2 (0.3%) | 1 (0.6%) |  |
| OR | 18 (2.4%) | 16 (2.7%) | 2 (1.3%) |  |
| Other | 2 (0.3%) | 2 (0.3%) | 0 (0%) |  |
| Out-of-Hospital | 40 (5.3%) | 36 (6.0%) | 4 (2.5%) |  |
| Hours on ECMO | 54 (24, 102) | 54 (23, 103) | 62 (29, 97) | 0.224 |
| Temperature Management |  |  |  | 0.036 |
| Missing | 36 (4.8%) | 33 (5.5%) | 3 (1.9%) |  |
| None | 46 (6.1%) | 34 (5.7%) | 12 (7.6%) |  |
| Targeted Hypothermia | 423 (56%) | 344 (57%) | 79 (50%) |  |
| Targeted Normothermia | 252 (33%) | 188 (31%) | 64 (41%) |  |
| Survival to Hospital Discharge | 200 (26%) | 162 (27%) | 38 (24%) | 0.448 |

**Table S3.**
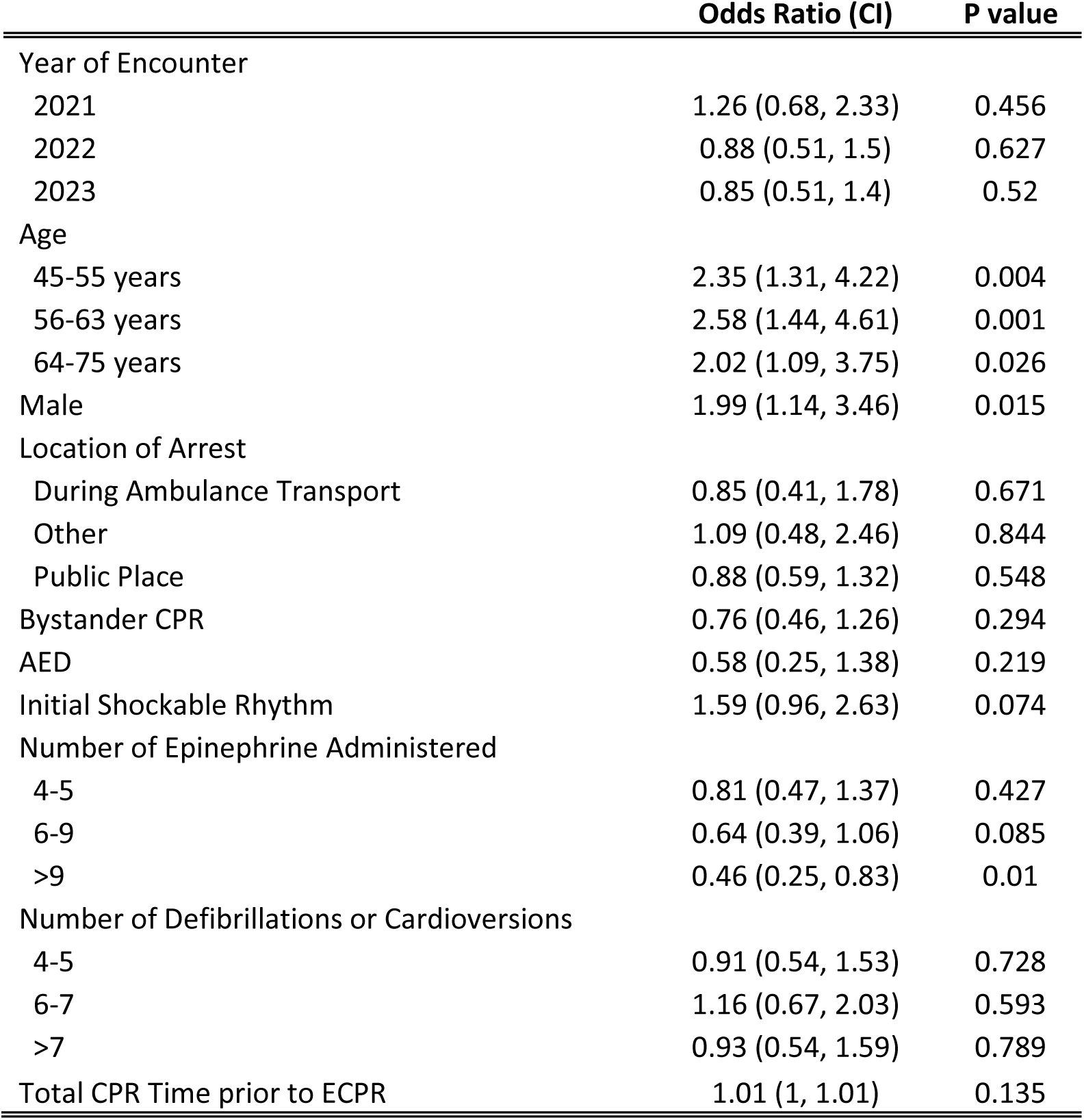
Predictors of percutaneous coronary intervention for the sensitivity analysis.

